# Efficacy of Ultrasound-Guided Erector Spinae Plane Block for Perioperative Pain Control and Short-Term Outcomes in Lumbar Laminoplasty

**DOI:** 10.1101/2020.01.30.20019745

**Authors:** Yanwu Jin, Shanshan Zhao, Jiahui Cai, Marcelle Blessing, Yongtao Sun, Shuai Hu, Qi Han, Xin Zhao, Haizhu Tan, Jinlei Li

## Abstract

**Background:** Erector spinae plane (ESP) block has been reported to provide analgesia in spine surgery in case reports or case series, and there have been no controlled studies to date evaluating its efficacy. We aimed to exam the roles of ESP block in lumbar surgery in a single center randomized control trial by injecting local analgesic into the interfacial plane between the erector spinae muscles and the transverse process under ultrasound guidance.

**Methods:** Consecutive elective lumbar surgery patients were randomized into either a control group (general anesthesia only, Group G, N=32) or a treatment group (general anesthesia plus ESP block, Group E, N=30). Several parameters including visual analog scale (VAS, primary outcome), perioperative anesthetics and analgesics usage, indexes of hemodynamics variation, return of bowel function and overall benefit of analgesia score (OBAS) were measured.

**Results:** Significant differences in VAS scores over time were found between the two matched groups (P = 0.010). Group E patients had significantly lower pain scores than Group G patients in the early postoperative period within the first 6 hr. Group G and Group E VAS scores peaked at 1 hr and 12 hr, respectively, and the peak pain score in Group G is significantly higher than that of Group E (P = 0.002). In addition, patients who received ESP block had lower perioperative analgesic and sedative medication requirements, improved satisfaction with pain management, more stable hemodynamics, and earlier bowel function return than those using general anesthesia alone.

**Conclusion:** Pre-incision bilateral single injection ESP blocks provided effective analgesia perioperatively during lumbar laminoplasty, decreased perioperative anesthesia and analgesia requirement, and accelerated short-term recovery.

## 1. INTRODUCTION

The concept of ESP block was first proposed by Mannion^1^ and further developed by Forero as an ultrasound-guided interfacial plane block, being successfully used in chronic thoracic neuropathic pain management^2^. ESP block provides analgesia by targeting the dorsal and ventral rami of the spinal nerves^2^. ESP blocks performed in the lumbar region for postoperative analgesia of lower extremity ^3-5^ and spine surgery^6 7^ have been described only in case reports and small case series. To date, no large-scale cohort studies or randomized controlled trials have been performed on its effectiveness for spine surgery. The aim of this study is to prospectively investigate the efficacy of the ESP block in pain management and short-term recovery after lumbar spine surgery.

## 2. MATERIALS AND METHODS

### 2.1 Study design

With institutional review board approval, this study is a single center, prospective, randomized, double-blind, controlled clinical trial conducted at a tertiary academic medical center. The study was retrospectively registered at http://www.chictr.org.cn on November 10, 2019, identifier ChiCTR1900026706.

### 2.2 Patients enrollment and randomization

A total of 72 consecutive patients who underwent elective lumbar laminoplasty for the treatment of lumbar spinal canal stenosis were identified from September 1^st^, 2018 to May 31^th^, 2019, 2 were excluded due to patient refusal, 70 were enrolled and randomly allocated into two groups using computer-generated randomization numbers contained in sealed opaque envelopes, 8 patients lost follow-up, and 62 patients were included in the statistical analysis (Figure 1). Inclusion criteria: elective laminoplasty and American Society of Anesthesiologists (ASA) physical status I ∼ III. Exclusion criteria: patient refusal, body mass index (BMI) less than 18 or higher than 35 kg/m^2^, age less than 18 years old, pregnant, history of relevant allergy to related perioperative medications, previous lumbar spine surgery, existing contraindications to nerve block such as coagulopathy, local and systemic infection, and unable to operate a patient-controlled analgesia (PCA) pump. All patients provided a written informed consent.

**Figure 1.**
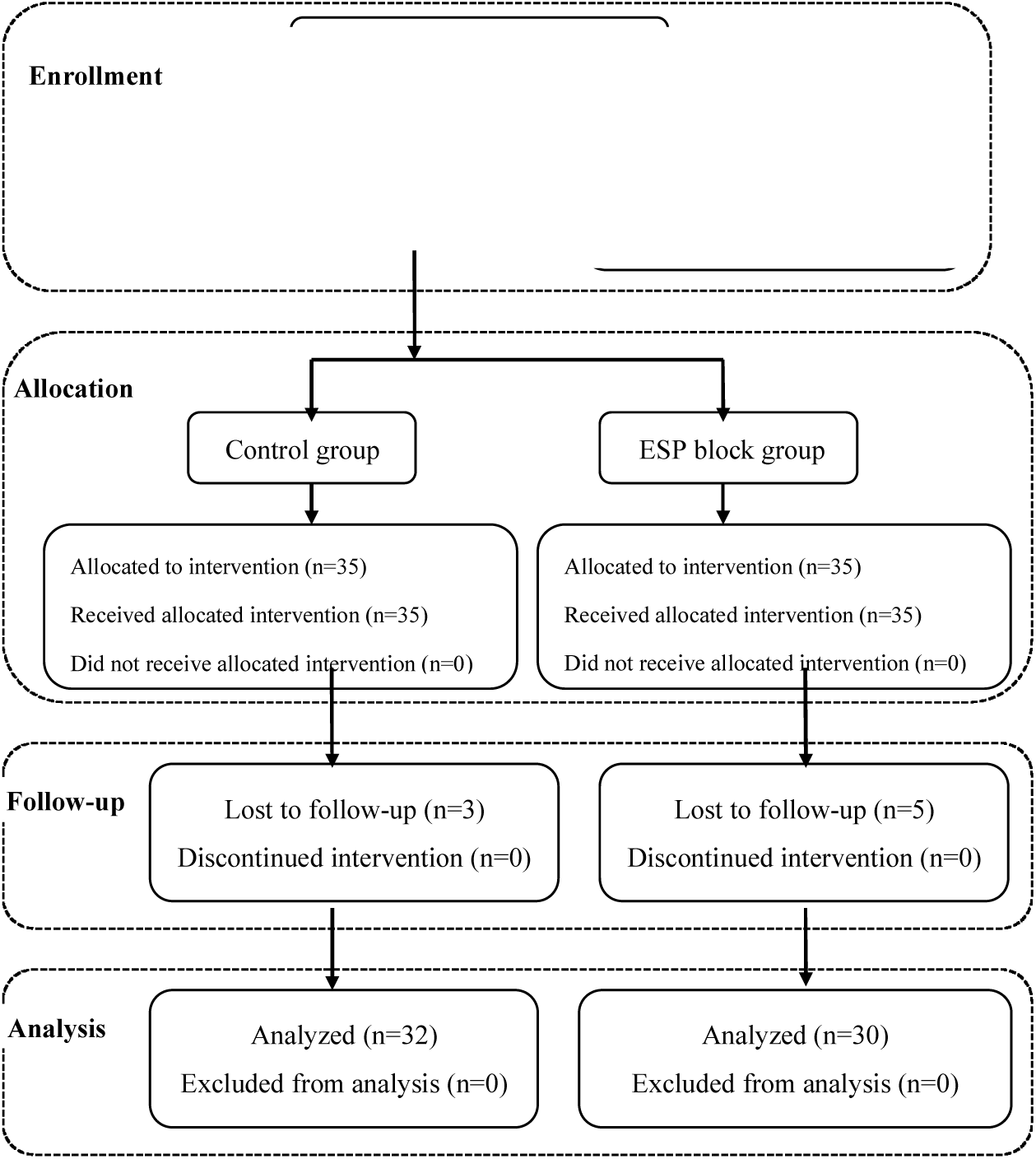
Flow Diagram in CONSORT Format.

The general anesthesia and subsequent ESP block under general anesthesia in prone position were performed by the same anesthesiologist. All patients were unaware of the grouping. Postoperative follow-up was performed by two certified nurses who were blinded to patient grouping.

### 2.3 Anesthesia and Analgesia

After American Society of Anesthesiologists (ASA) standard monitors were applied, an awake radial arterial line was placed, induction of general anesthesia was standardized in the two groups using propofol (1.5 ∼ 2.5 mg/kg) and sufentanil (0.2 ∼ 0.3 µg/kg). Endotracheal intubation was facilitated with cis-atracurium (0.2 mg/kg) and maintenance of anesthesia and analgesia was achieved with sevoflurane, intermittent sufentanil and cis-atracurium. About 30 minutes before the end of the surgery, remifentanil infusion was started to facilitate extubation. Group G received general anesthesia only, while Group E received general anesthesia plus pre-incision ESP blocks with 20 ml 0.375% ropivacaine hydrochloride injected on each side.

All patients were admitted to the surgical intensive care unit for 48 hours after the surgery. The primary outcome was the VAS pain scale for the first 48 hours after the surgery. The secondary outcomes included intraoperative sufentanil and remifentanil consumption, postoperative sufentanil consumption for the first 48 hours, pain management satisfaction indicated by OBAS score, intraoperative sevoflurane consumption, number of postoperative PCA attempts, postoperative rescue analgesics usage, adverse events such as nausea, vomiting, dizziness and irritability, and functional outcomes such as return of bowel function indicated by first anal exhaust time, length of hospitalization, and hospitalization expenses.

All patients were provided with a PCA pump with a mixture of sufentanil 1µg/ml, dezocine 0.2 mg/ml, dexmedetomidine 1 µg/ml, and palonosetron 0.25 mg at the basal rate 2 ml/hour, a 2 ml per demand dose with a lock-out time of 15 minutes. Intravenous sodium parecoxib and intramuscular pethidine are the first and second line treatment for breakthrough pain respectively. Nausea and vomiting were managed with intravenous palonosetron as needed.

### 2.4 Ultrasound-guided ESP block

The ESP block was performed after induction of general anesthesia with patients in prone position. A 1.4-5.1 MHz low-frequency probe (C5-1s convex transducer, Mindray M8 Super, Shenzhen, China) was first placed in a longitudinal orientation in the midline to identify the spinous process of at the appropriate lumbar level, then scanned laterally about 2-3 centimeters until visualization of the paraspinal muscles and the transverse process at the same time, as shown in Figure 2b. After standard sterilization, a 21-G short bevel nerve block needle (PAJUNK Gmbh Medizintechnologie, Geisingen, Germany) was advanced in a cephalad-to-caudad direction, in-plane under real-time ultrasound guidance, through skin, subcutaneous tissue and erector spinae muscles until reaching the transverse process. After negative aspiration of blood or cerebral spinal fluid, a small volume of local anesthetic was injected to confirm the position of the needle tip between the erector spinae muscles and the transverse process. A total of 20 ml of 0.375% ropivacaine was incrementally injected with intermittent negative aspiration on each side.

**Figure 2a.**
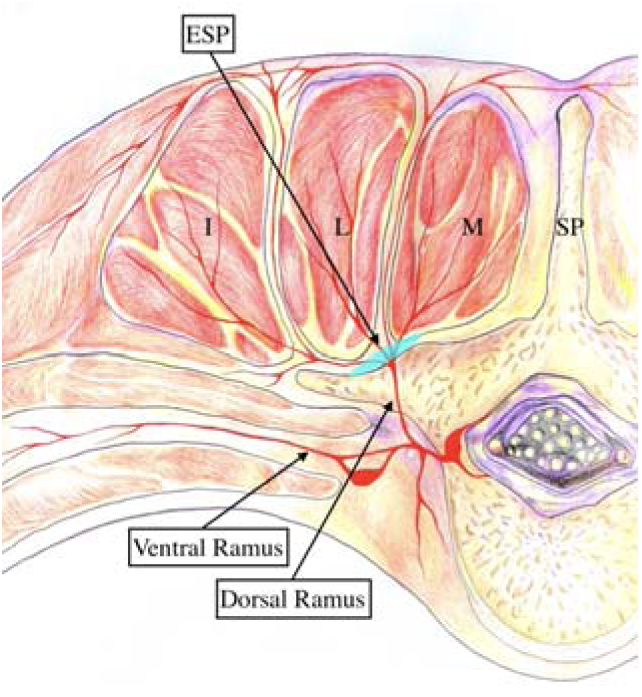
Illustration of Erector Spinae Block. I: Iliocostalis muscle, L: Longissimus muscle, M: Multifidus muscle, SP: Spinous process, ESP: Erector spinae plane

**Figure 2b.**
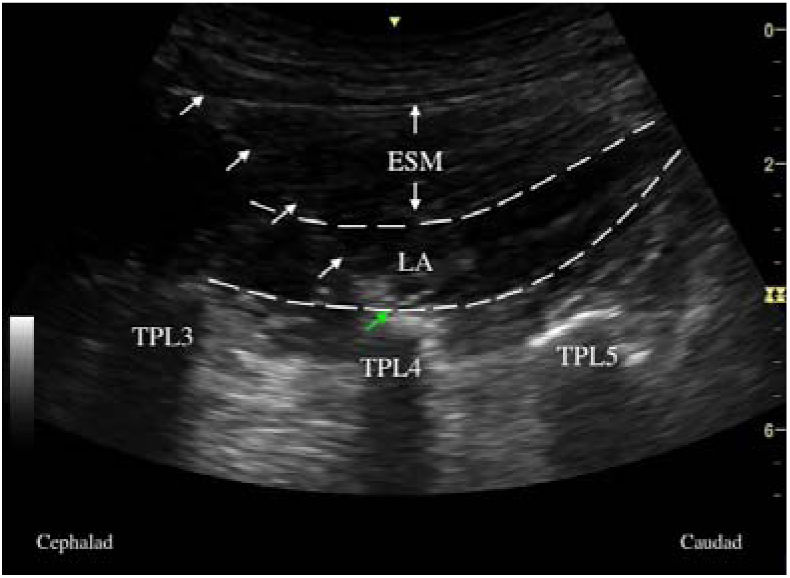
Sonographic Illustration of Erector Spinae Plane (ESP) Block. ESM: Erector spinae muscle; LA: Local anesthetic; TPL, Lumbar transverse Process; White arrow: Needle trajectory; Green arrow: Needle tip

### 2.5 Statistical Analysis

The sample size calculation and all statistical data analysis was performed with R software. The significance level was set at a P value <0.05.

To estimate the sample size and power, 20 patients were randomly assigned into two groups in the preliminary pilot study. For primary outcome (VAS pain scores for the first 48 hours), we first applied the sample size formula for two-sample longitudinal data to compute the predicted power using two-sided test. Possible new enrolled sample size would be obtained. We repeated this procedure until the predicted power achieved the preset power β (β = 0.8). For secondary outcomes (analgesics consumption), we again calculated the sample size and power under the hypothesis tested. Combining all predicted sample size together, the estimated sample size was about 28 for each group. Considering 10% losses of follow-ups, the effective sample size was set at 36 for each group.

Data are presented as mean (SD), or median (interquartile range (IQR): 25th percentile - 75th percentile) for continuous variables, and frequency and percentage for categorical variables. The study for our primary outcome, VAS at 1h, 3h, 6h, 12h, 24h, and 48h after operation, was a repeated measures design because multiple measurements were on the same unit and observed over time^8^. However, VAS over times tended to correlate with each other. It was therefore important to account for the correlation between measurements. We applied Mauchly’s Test of Sphericity to test the correlations between measurements over times^9^.

In equation (1),

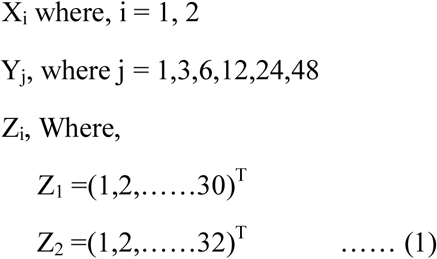

We let Y_j_ be VAS over time, X_i_ be types of anesthesia, Z be patients in each group. i be the number of group,and j be time interval.

From equation (1), we found that multivariate analysis of variance (MANOVA) couldn’t be applied to analyze the difference of Y_j_, between two groups. Ball Divergence Test (BD)^10^ is a nonparametric two samples test. It is a measure of the difference between two probability measures in separable Banach spaces^10^. Hence, BD was adopted to identify the difference for VAS over time between the two groups. In this study, let X_i_ and Y_j_ to express two groups’ patients measurements of VAS overtimes.

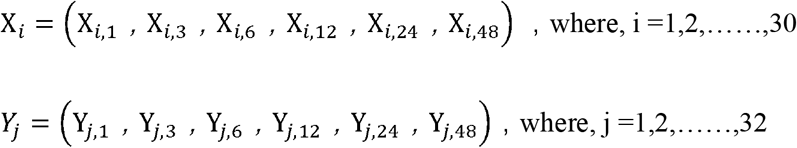

Defined the distance between *X*_*i*_’s and γ_*j*_’s as the equitation (2)

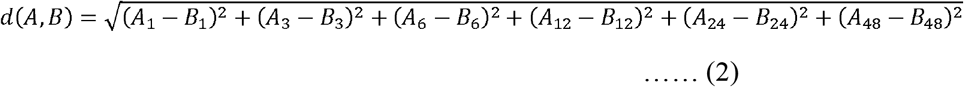

We used equitation (2) to calculate the distance in our study as:

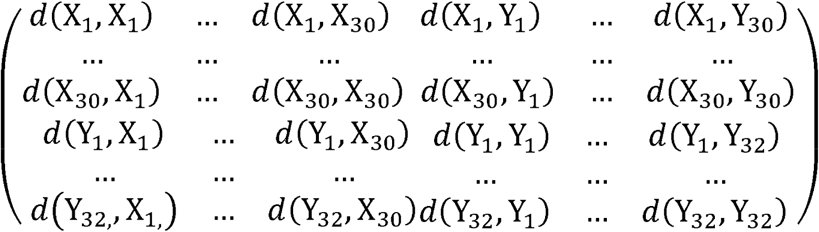

We subsequently compute the test statistic with techniques described by Zhang^10^.

After comparing the VAS difference between the two groups using Ball divergence test, we subsequently studied the change pattern as well as the extreme points over time between the groups.

Two samples rank sum test was applied to analyze the cardinal variables with non-normal distribution (such as, intraoperative sevoflurane consumption, intraoperative inhalation concentration of sevoflurane) or ordinal scale variables (such as ASA status, New York Heart Association (NYHA) status). Two-sample t test was used for data with normal distributions such as postoperative sufentanil consumption, intraoperative remifentanil consumption, and first anal exhaust time. The two samples test for binomial proportions or Fisher exact test were used to analyze the difference between the two groups on the number of spine level involved during the surgery, postoperative nausea/vomiting. All tests were one-sided test.

## 3. RESULTS

### 3.1 Baseline Characteristics

There was no significant difference between groups in major demographic variables, such as age, gender, BMI, history of opioid usage, and ASA status. In addition, there were no significant difference in the number of lumbar spine segment involved (P= 0.296), surgical time (P= 0.136) and general anesthesia duration (P= 0.136) (Table 1).

**Table 1.**
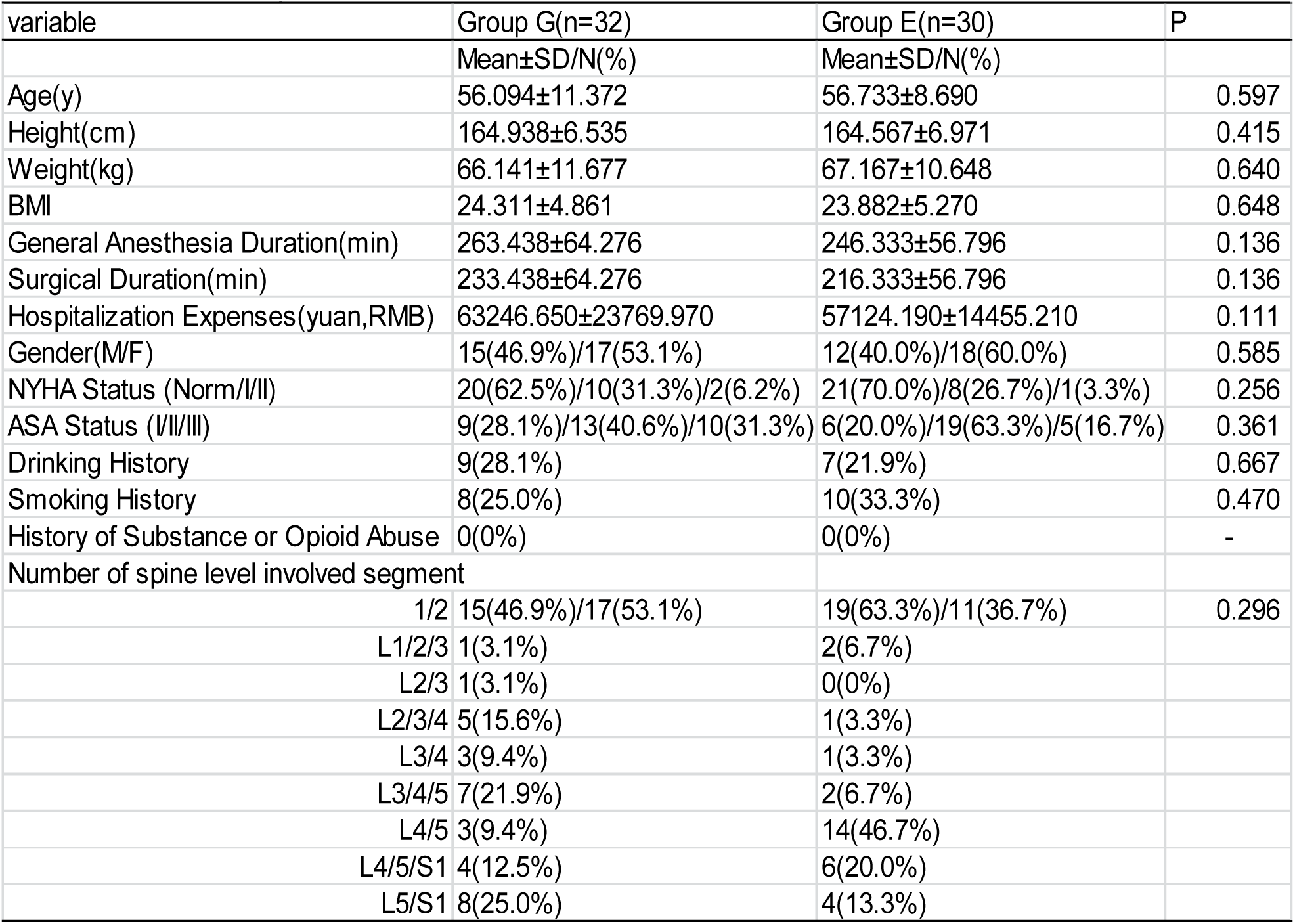
Patients demographics

### 3.2 Primary outcome: VAS pain scores for the first 48 hours

The result of Mauchly’s Test of Sphericity showed that the correlations between each two neighboring time points (P= 0.000) were strong. BD result indicated that there were significant differences in VAS scores over time between the two matched groups (P= 0.010). From the time line chart (Figure 3), patients in Group E reported less pain than patients in Group G in the early postoperative period, postoperative VAS at 1 hr, 3 hr, 6 hr in Group G and Group E were 4.250±1.414 vs 2.033±0.718, P=0.000, 3.906±1.14 vs 2.400±0.814, P= 0.000, and 3.656±1.066 vs 2.767±0.679, P=0.000 respectively. Analysis of peak pain scores in Group G and Group E at 1 hr and 12 hr respectively indicated that Group G patients experienced greater pain than that of Group E (P= 0.002). In addition, the OBAS pain satisfaction score of Group E was significantly lower than that of Group G (P=0.008), where lower OBAS score reflects more benefit a patient gained from a specific pain management (Table 2).

**Table 2.** Intraoperative Hemodynamics and Perioperative Anesthetics/Analgesics

|  |  |  |  |
| --- | --- | --- | --- |
| Variable | Group G (n=32) | Group E (n=30) | P-value |
|  | Mean±SD | Mean±SD |  |
| hemodynamics |  |  |  |
| □SBP (mmHg) | 8.000±4.500 | 7.500±4.000 | 0.177 |
| □DBP (mmHg) | 6.000±4.000 | 3.5000±3.000 | 0.000 |
| □HR (bpm) | 3.000±3.000 | 1.500±2.750 | 0.003 |
| Postoperative pain score |  |  |  |
| Postoperative VAS Score at 1 Postoperative Hour (0-10) | 4.250±1.414 | 2.033±0.718 | 0.000 |
| Postoperative VAS Score at 3 Postoperative Hours (0-10) | 3.906±1.146 | 2.400±0.814 | 0.000 |
| Postoperative VAS Score at 6 Postoperative Hours (0-10) | 3.656±1.066 | 2.767±0.679 | 0.000 |
| Postoperative VAS Score at 12 Postoperative Hours (0-10) | 3.281±0.958 | 3.300±0.877 | 0.629 |
| Postoperative VAS Score at 24 Postoperative Hours (0-10) | 2.656±0.701 | 2.600±0.724 | 0.424 |
| Postoperative VAS Score at 48 Postoperative Hour (0-10) | 1.969±0.595 | 1.867±0.571 | 0.250 |
| anesthetics and analgesics consumption |  |  |  |
| OBAS at 24 postoperative hours | 3.000±2.000 | 2.000±1.000 | 0.008 |
| Intraoperative inhalation concentration of sevoflurane | 2.500±0.625 | 1.500±0.500 | 0.000 |
| Intraoperative Sevoflurane Consumption (ml) | 79.200±23.625 | 46.800±17.100 | 0.000 |
| Intraoperative Sufentanil Consumption (μg) | 45.000±1.250 | 25.000±5.000 | 0.000 |
| Postoperative Sufentanil Consumption (μg) | 75.375±9.349 | 65.067±13.421 | 0.000 |
| Intraoperative Remifentanil Consumption (μg) | 188.531±52.652 | 96.400±27.399 | 0.000 |
| The number of PCA Attempts | 2.500±2.000 | 2.000±3.750 | 0.037 |
| Number of Rescue Analgesia Received | 1.000±2.000 | 1.000±1.000 | 0.215 |
| Number of Patient Using Pethidine | 0 | 0 | - |

**Figure 3.**
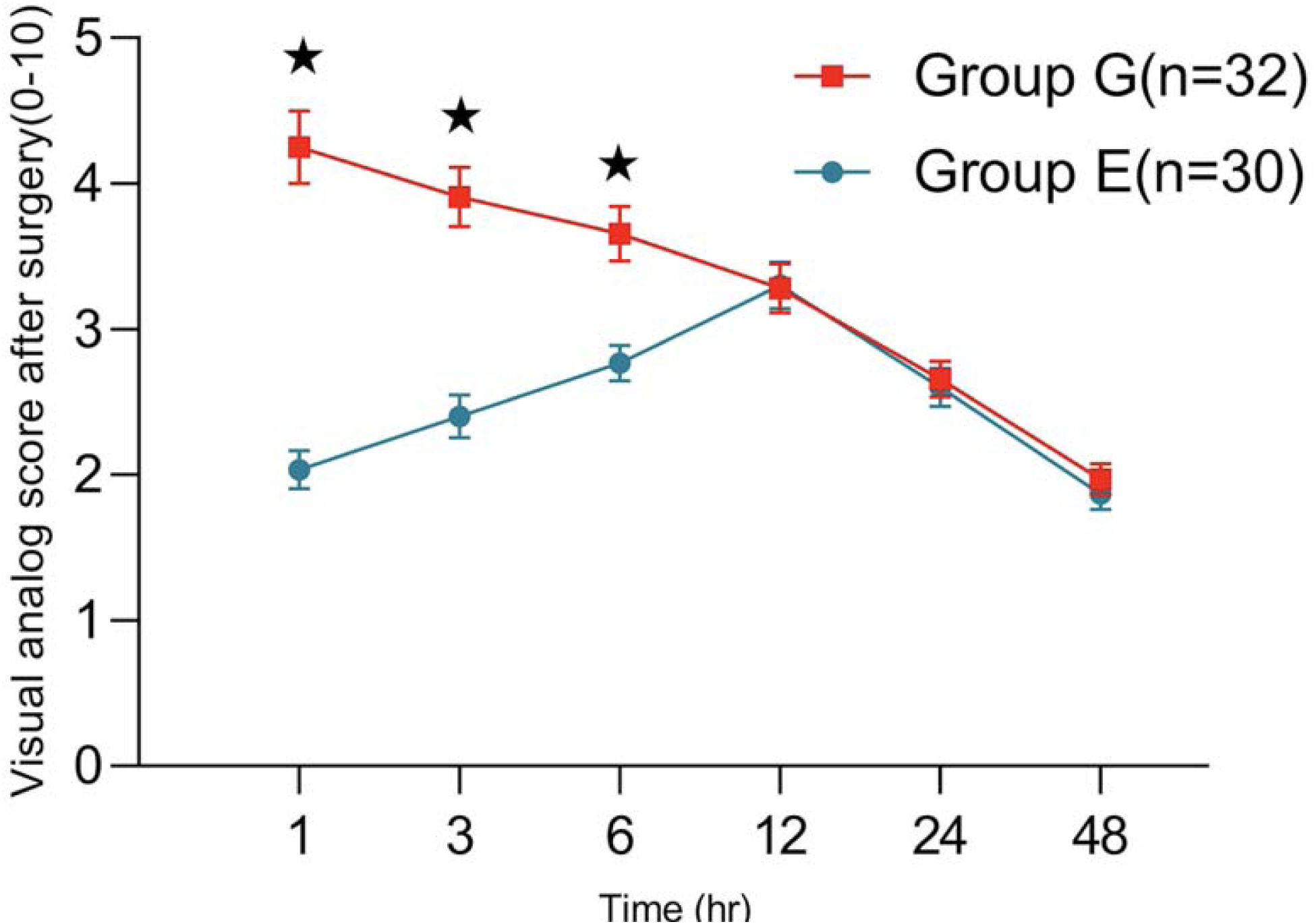
VAS Pain Score Time Line Chart. ^⍰^P < 0.05. Data are presented as mean ± SD.

### 3.3 Secondary outcomes: hemodynamics, analgesics consumption and recovery

Intraoperative hemodynamics was monitored continuously through a radial arterial line and there was no significant difference of systolic blood pressure changes (⍰SBP) one minute after incision between the two groups (P= 0.177), but the diastolic blood pressure change (⍰DBP) and heart rate change (⍰HR) in Group G were higher than that of Group E (P_⍰DBP_ = 0.000, P_⍰ HR_ = 0.003, Table 2).

The anesthetics and analgesics consumption of Group G were significantly higher than that of Group E (Table 2), including intraoperative inhaled concentration of sevoflurane (P=0.000), intraoperative sevoflurane consumption (P = 0.000), intraoperative remifentanil consumption (P = 0.000), intraoperative sufentanil consumption (P = 0.000), and postoperative sufentanil consumption (P = 0.000).

The first anal exhaust time of patients in Group E was earlier than that of Group G (P = 0.014). There was no difference in the cost of hospitalization (P = 0.562) or length of hospital stay (P= 0.111) (Table 3)

**Table 3.** Postoperative Recovery

| Table 3. Postoperative Recovery |  |  |  |
| --- | --- | --- | --- |
|  | Group G (n=32) | Group E (n=30) | P-value |
|  | Mean±SD/N(%) | Mean±SD/N(%) |  |
| Hospital Stay(hr) | 395.344±77.994 | 398.767±92.562 | 0.562 |
| Hospitalization Expenses(yuan,RMB) | 63246.650±23769.970 | 57124.190±14455.210 | 0.111 |
| First Anal Exhaust Time (hr) | 39.875±11.494 | 32.000±15.682 | 0.014 |
| Postoperative nausea | 11(34.4%) | 6(5.0%) | 0.205 |
| Postoperative vomiting | 5(15.6%) | 4(13.3%) | 1.000 |
| Postoperative Dizziness | 1(3.1%) | 0(0%) | - |
| Postoperative Irritability | 0(0%) | 0(0%) | - |

## 4. DISCUSSION

Degenerative disk disease is a common orthopedic condition in clinical practice. Spine surgery in the thoracolumbar region is frequently performed to relieve pain and disability secondary to spinal stenosis^11^. During the surgery, the paraspinal muscles are dissected away from the spinous process and lamina to the lateral side of the facet joint. Mechanical and thermal trauma during surgery may cause muscle ischemia and damage to nerves innervating the paraspinal muscles. Severe postoperative incisional pain as well as pain to the surrounding soft tissue and bone is common. Opioid agonists administration slows down normal gastrointestinal motility, increases the incidence of respiratory complications, and prolongs hospital stay^12^. Enhanced recovery after surgery protocols frequently include the use of regional anesthesia techniques to minimize opioid analgesics whenever possible^13 14^. Although using regional anesthesia to control postoperative pain has been shown to have morbidity and mortality benefits, regional anesthesia options have been limited for spine surgery^12 13 15-17^.

There are several case reports and case series on the effects of peripheral nerve block such as thoracolumbar interfascial plane (TLIP)^18 19^ or ESP^6 7^ block in spine surgeries. To our best knowledge, this study is the first of a relatively large sample size and to be prospectively performed with a randomized and controlled design. Our study showed that the ESP block provided improved analgesia with significantly lower VAS scores at rest at 1, 3, and 6 hr postoperatively, decreased intraoperative and postoperative opioid consumption and less intraoperative sevoflurane requirements. OBAS, a validated multi-dimensional tool to assess pain intensity, opioid-related adverse effect and patient satisfaction of pain control, was also shown to be better in Group E as compared to group G. Finally, effective analgesia via opioid-reducing ESP block placed preoperatively was shown to accelerate bowel function recovery postoperatively.

There are several limitations in this study. Anatomically we could not measure the exact local anesthetic spread after injection and did not assess the dermatomal coverage post block placement either. Secondly, VAS scores with activity were not assessed. In addition, this study focused on short-term outcomes for the first 48 hours after surgery, and long-term outcomes such as the efficacy of the ESP block in persistent postsurgical pain prevention or long-term surgical outcome remains unknown. A larger scale study, ideally a multicenter, prospective study, may better address these questions.

## 5. Conclusions

The ultrasound guided ESP block can provide effective analgesia, decrease perioperative opioid consumption and is beneficial to the recovery of gastrointestinal function in patients undergoing lumbar laminoplasty.

## Data Availability

The study was registered at http://www.chictr.org.cn, identifier ChiCTR1900026706.

## ACKNOWLEDGEMENTS

None.

